# Modeling the impact of COVID-19 vaccination in Lebanon: A call to speed-up vaccine roll out

**DOI:** 10.1101/2021.05.27.21257937

**Authors:** Ghina R. Mumtaz, Fadi El-Jardali, Mathilda Jabbour, Aya Harb, Laith J. Abu-Raddad, Monia Makhoul

## Abstract

**Background:** Amidst a very difficult economic and political situation, and after a large first SARS-CoV-2 wave near the end of 2020, Lebanon launched its vaccination campaign on 14 February 2021. To date, only 6.7% of the population have received at least one dose of the vaccine, raising serious concerns over the speed of vaccine roll-out and its impact in the event of a future surge.

**Objective:** Using mathematical modeling, we assessed the short-term impact (by end of 2021) of various vaccine roll-out scenarios on SARS-CoV-2 epidemic course in Lebanon.

**Results:** At current immunity levels in the population, estimated by the model at 40% on 15 April 2021, a large epidemic wave is predicted if all social distancing restrictions are gradually eased and variants of concern are introduced. Reaching 80% vaccine coverage by end of 2021 will flatten the epidemic curve and will result in a 37% and 34% decrease in the peak daily numbers of severe/critical disease cases and deaths, respectively; while reaching intermediate coverage of 40% will result in only 10-11% decrease in each. Reaching 80% coverage by end of 2021 will avert 3 times more hospitalizations and deaths over the course of this year compared with 40% coverage. Impact of vaccination was substantially enhanced with rapid scale-up. Reaching 80% vaccine coverage by August would prevent twice as many severe/critical disease cases and deaths than if it were reached by December. Finally, a longer duration over which restrictions are eased resulted in a more favorable impact of vaccination.

**Conclusion:** For vaccination to have an impact on the predicted epidemic course and associated disease burden in Lebanon, vaccination has to be rapid and reach high coverage (at least 70%), while sustaining social distancing measures during roll-out. At current vaccination pace, this is unlikely to be achieved. Concerted efforts need to be put to overcome local challenges and substantially scale up vaccination to avoid a surge that the country, with its multiple crises and limited health-care capacity, is largely unprepared for.

## Introduction

Lebanon documented its first severe acute respiratory syndrome coronavirus 2 (SARS-CoV-2) case on February 21, 2020. After almost a year of ongoing cycles of imposing and easing restrictions to suppress the epidemic, the country experienced its first large wave in December 2020-January 2021 which stretched its fragmented, under-resourced, and highly privatized health care system beyond capacity. As of May 19, 2021, over 537,000 confirmed cases and 7,650 deaths have been documented [1], for a population of 6.8 million inhabitants [2].

While the pandemic has severely impacted economies and societies worldwide, it was compounded in Lebanon by multiple crises including an unpreceded devaluation of the local currency, political uprising against the ruling class, and the world’s largest non-nuclear explosion that blasted its capital Beirut in August 2020 causing thousands of casualties and destroying several hospitals with Coronavirus Disease 2019 (COVID-19) units. With over one million refugees on its territory, Lebanon also hosts the highest per capita number of refugees worldwide [3], a population highly vulnerable to large SARS-CoV-2 outbreaks [4]. At this stage, mass vaccination with the recently available and highly efficacious COVID-19 vaccines [5-9] brings hope to control infection spread and mitigate its profound impact, especially with the emerging ‘real-world’ evidence of their high effectiveness in reducing both symptomatic and asymptomatic disease [10-13].

On February 14, 2021 the Lebanese Ministry of Public Health launched its vaccination campaign and inoculated its first residents. To date, doses enough to ensure a 73% coverage of the population have been reserved. However, only 7% of these doses have been received so far and, three months into the vaccination campaign, only 6.7% of Lebanese residents have received at least one dose of the vaccine [14], raising serious concerns over supply and the speed of vaccine roll-out. Also, only 21% of the population have registered to receive the vaccine on the ministry’s official vaccine platform [14], raising further concerns over demand, access to and use of the online registration platform, and misinformation among the general public.

With the expected relaxation of social distancing restrictions in the upcoming summer months and with the threat of the potential introduction of more transmissible and likely more severe SARS-CoV-2 variants [15-17], the objective of this study was to forecast the short term (by end of year) impact of COVID-19 vaccination in Lebanon in order to inform policy and the national vaccination strategy. We simulate what would be the epidemic course if all restrictions are gradually eased and if variants of concern are introduced, and assess the impact of different levels of vaccine coverage, different durations for vaccine scale-up, and different schedules for easing social distancing restrictions on SARS-CoV-2 incidence and associated burden of COVID-19 disease and death.

## Methods

### Mathematical model description

We adapted an existing mathematical model that investigates the generic population-level impact of SARS-CoV-2 vaccination and extended it to be applied to Lebanon [18-20]. This is a deterministic age-structured Susceptible-Exposed-Infectious-Recovered (SEIR) model that stratifies the population into compartments reflecting their vaccination status, age group, infection status, infection stage, and disease stage. The model consists of a set of coupled nonlinear differential equations that depict population movement between the different compartments. An age-mixing matrix allowed a range of assortativeness for contact patterns between individuals in different age groups. Population demography was assumed to be stable given the short-term timeframe of the predictions (one year). We assumed that the duration of natural immunity (following recovery from a natural infection) and vaccine-acquired immunity is one year [21-25]. We further assumed that waning of immunity follows a gamma distribution where only a small fraction of individuals lose their immunity either much earlier or much later than one year. The model was coded, fitted, and analyzed using MATLAB R2019a [26]. Details of the model structure and equations can be found in the Supplementary Information Text S1A, S1B, and Table S1.

### Model parameterization, fitting, and application to Lebanon

The model was parametrized with most recent SARS-CoV-2 epidemiological and natural history data. Model parameters, values, and justifications can be found in the Supplementary Information Text S1C and Table S2. The model was fitted to SARS-CoV-2 case series of infections and deaths in Lebanon [1]. Fitting used a nonlinear least square technique, based on the Nelder-Mead simplex algorithm. Given the large wave that happened near the end of year holidays and informed by unpublished clinical antibody testing reports, we assumed that 20% of the Lebanese population had already been exposed to the infection by January 1, 2021.

Since the ultimate objective of vaccination is to allow a return of social and economic life back to normalcy, we gradually eased all social and physical distancing restrictions over a period of four months starting 15 April 2021. We assumed a constant SARS-CoV-2 incidence rate between 1 January 2021 and 15 April 2021, to reflect the observed relative stability of the epidemic during this time where an average of 3,000 confirmed daily cases were being reported [1]. As restrictions were gradually eased starting 15 April 2021, R_0_ was gradually increased to reach R_0_=6 at the end of the four months, thus accounting for the increased transmissibility in absence of non-therapeutic interventions of the new SARS-CoV-2 variants of concern [15, 27-31].

### Vaccine characteristics and scale-up scenarios

In Lebanon, the BNT162b2 mRNA Covid-19 Vaccine (Pfizer) is the main vaccine rolled out and has the largest fraction of the doses reserved, followed more recently by the ChAdOx1 nCoV-19 vaccine (Astrazeneca). We assumed that the vaccine has ‘real-world’ effectiveness of 80% at reducing infection (VE_S_=80%), based on recent evidence for the effectiveness of the vaccine in reducing asymptomatic as well as symptomatic infections [10, 12, 32].

Vaccination was introduced on 14 February 2021 (actual date) and scaled-up at a linear rate to reach the desired coverage by 31 December 2021, as informed by the schedule of incoming vaccine shipments to Lebanon where more doses are expected to become available over time. Two scenarios are presented: 1) 80% vaccine coverage is reached by the end of the year (target coverage rate) and 2) 40% vaccine coverage is reached by the end of the year (more realistic scenario given current vaccination roll out). During vaccine roll-out, we assumed that, at each point in time, all individuals in the population have an equal chance of getting vaccinated, including those that had already acquired the infection naturally and recovered from it. Vaccine coverage was defined as the proportion of the population that had received two-doses of the vaccine and completed 14 days after the second dose at a given point in time.

### Measures of vaccine impact

After easing all restrictions over four months and introducing variants of concern starting 15 April 2021, we assessed the population-level impact of SARS-CoV-2 vaccination by quantifying and comparing the incident number of infections, severe/critical disease cases, and deaths for the scenarios of 1) no vaccination, 2) vaccine introduced on 14 February 2021 and 80% vaccine coverage is reached by the end of the year, and 3) vaccine introduced on 14 February 2021 and 40% vaccine coverage is reached by the end of the year. We used the World Health Organization (WHO) guidelines for classifying infection severity [33] and determining COVID-related death [34].

We further compared the cumulative number of averted severe/critical disease cases and deaths from 14 February 2021 (date of vaccine introduction) to 31 December 2021 for a range of vaccine coverage attained by end of year, for different durations of vaccine scale-up to reach the target 80% coverage (thus assessing impact of the speed of rollout), and for different durations over which restrictions are eased while reaching the target 80% coverage by end of year 2021. We did not assess the impact of vaccine prioritization by age as it was investigated in earlier studies [18, 35].

### Uncertainty analysis

Uncertainty analysis was conducted to investigate the impact of uncertainty around our assumed transmissibility of the new variants of concern introduced on 15 April 2021 on the model’s predictions of the COVID-19 severe/critical disease incidence for the different scenarios. For a total of 500 simulation runs, we used, in each run, Latin Hypercube sampling [36, 37] to select an R_0_ within ±30% of the assumed baseline value of R_0_=6. The resulting distribution for the predicted number of incident and cumulative severe/critical disease cases across the 500 runs was used to calculate the predicted mean and 95% uncertainty intervals around these two estimates.

## Results

Based on the model assumptions, we estimated that 44% of the Lebanese population have acquired either natural or vaccine-induced immunity to SARS-CoV-2 by 15 April 2021. With this herd immunity level in the population, gradually lifting all social distancing restrictions by mid-August 2021 and introducing more transmissible variants of the virus will result in a large new epidemic wave over the summer months (Figure 1). While it did not fully prevent the new surge, the 80% vaccination coverage flattened the epidemic curve (yellow curve), resulting in a smaller epidemic. Under this scenario, the peak incidence of infections, severe/critical disease cases, and deaths are reduced by 37.1%, 36.6%, and 34.2%, respectively, compared with the no vaccination scenario (Figure 1).

**Figure 1.**
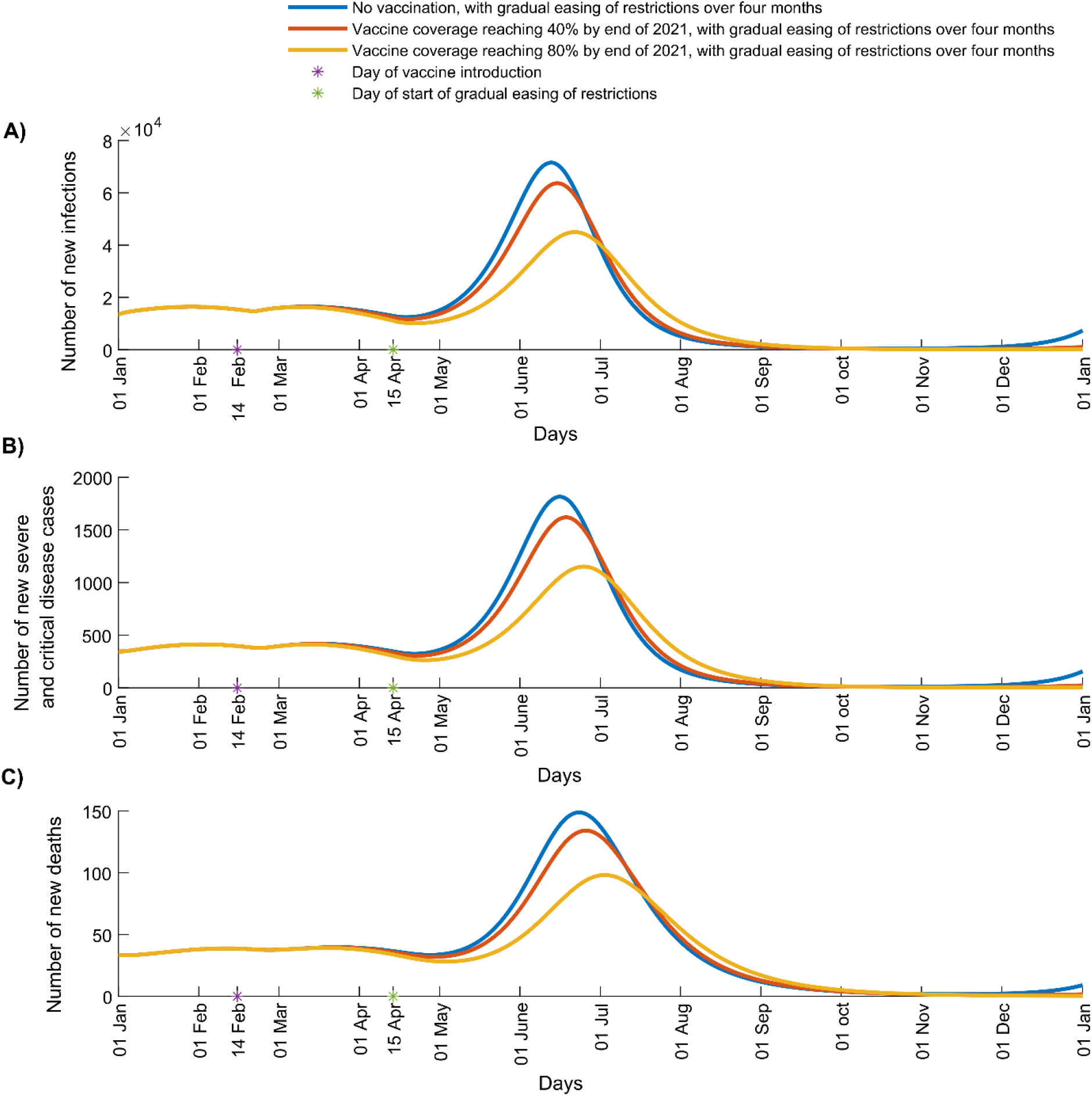
Impact of SARS-CoV-2 vaccination on number of A) new infections, B) new severe and critical disease cases, and C) new deaths in Lebanon. The vaccine is introduced on 14 February 2021 and vaccine coverage is scaled up to reach 40% (red curve) or 80% (yellow curve) by 31 December 2021. The simulations assume R_0_ of 1.2 from 1 January 2021 to April 15 2021 when it starts to increase with gradual easing of restrictions to reach 6.0 after four months.

Vaccination did not fully prevent a new epidemic surge because the current contact rate in the community is not sufficiently low and the population immunity level is not sufficiently high, and because of the higher transmissibility of the new variants.

The 40% vaccination coverage was not able to noticeably flatten the curve (Figure 1, orange curve) and hence had a much smaller impact on epidemic course compared with the target 80% coverage rate. This scenario resulted only in an 11.0%, 10.8%, and 10.0% decrease in the peak incidence of infections, severe/critical cases, and deaths, respectively, compared with the no vaccination scenario (Figure 1). Reaching 40% coverage by the end of the year means that most vaccinations would be administered after the predicted time of the summer surge, which undermined vaccine impact under this scenario.

The cumulative numbers of averted severe/critical disease cases and deaths increased substantially and steadily with higher vaccine coverage at the end of 2021 (Figure 2). For example, reaching 80% vaccine coverage will result in a 3-fold increase in the number of prevented severe/critical cases and deaths compared with 40% vaccine coverage rate. The gains in averted disease cases and deaths increased sharply as vaccine coverage reaches 70%. Reaching 80% vaccination coverage by end of the year will prevent a total of 23,600 cumulative severe/critical cases and 2,188 cumulative deaths between 14 February 2021 and 31 December 2021 compared with the no vaccination scenario. (Figure 2).

**Figure 2.**
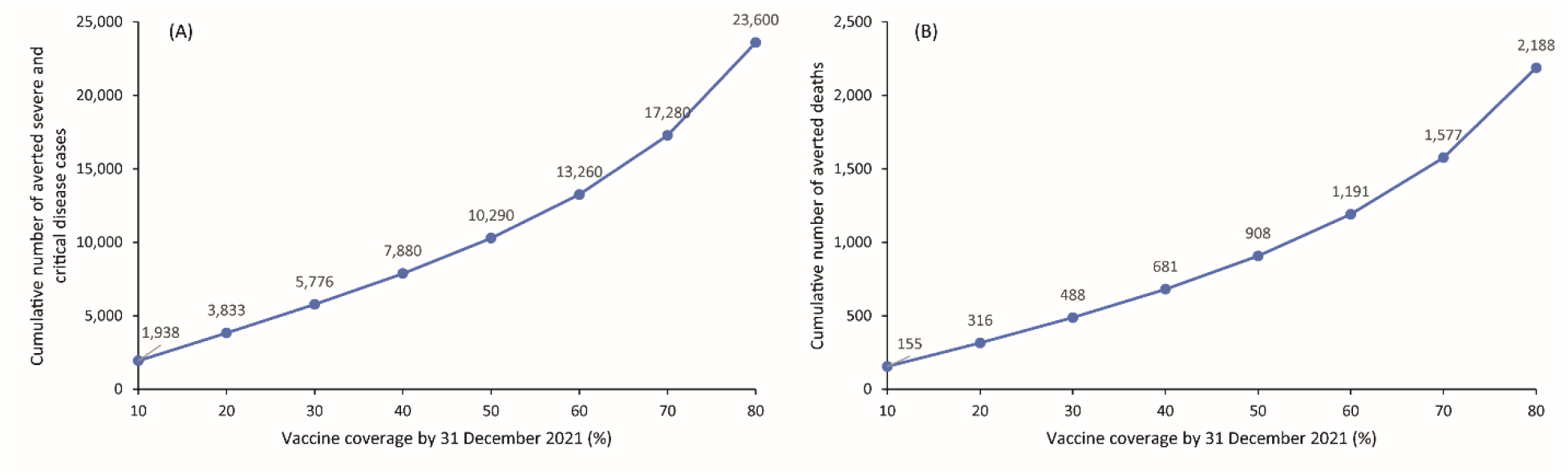
Impact of various levels of vaccine coverage reached by 31 December 2021 on the cumulative number of averted (A) severe and critical disease cases and (B) deaths from 14 February 2021 (date of vaccine introduction) to 31 December 2021. The scenarios assume gradual easing of all restrictions over four months starting 15 April 2021.

The effectiveness of the vaccine in preventing severe/critical disease and death was substantially enhanced by more rapid scale-up to reach the 80% target vaccine coverage by end of the year (Figure 3). For example, reaching 80% vaccine coverage by August would prevent twice as many severe/critical disease cases and deaths than if it were reached by December (Figure 3).

**Figure 3.**
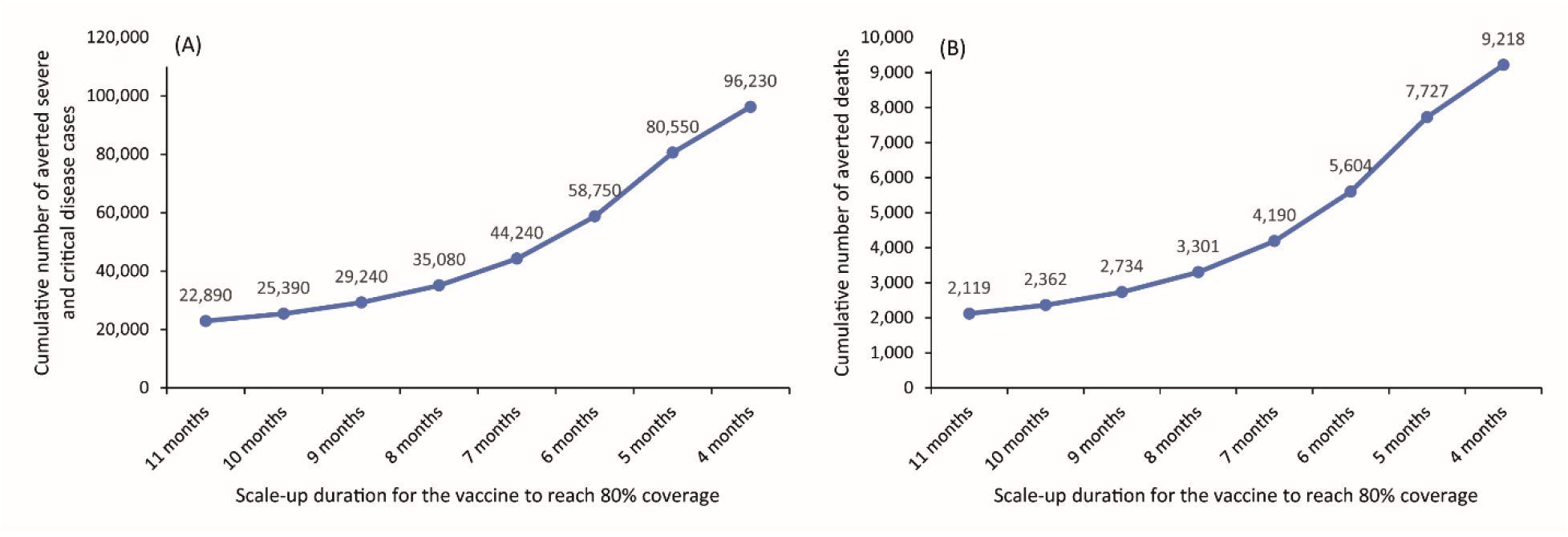
Impact of vaccine scale-up duration to reach 80% coverage by 31 December 2021 on the cumulative number of averted (A) severe and critical disease cases and (B) deaths from 14 February 2021 (date of vaccine introduction) to December 31, 2021. The scenarios assume gradual easing of all restrictions over four months starting 15 April 2021.

In all of the above scenarios, we assume that easing of all restrictions would occur over four months starting on April 15, 2021. However, as expected, we found that a longer duration over which restrictions are eased would result in a more favorable impact of vaccination (Figure 4). For example, if easing of restrictions happens over 8 months, that is people would still exert some levels of social distancing until December 2021 when the target 80% vaccine coverage is reached, twice as many deaths would be averted than if social contacts are back to normalcy by August 2021 (4,175 versus 2,185 averted deaths, respectively) (Figure 4).

**Figure 4.**
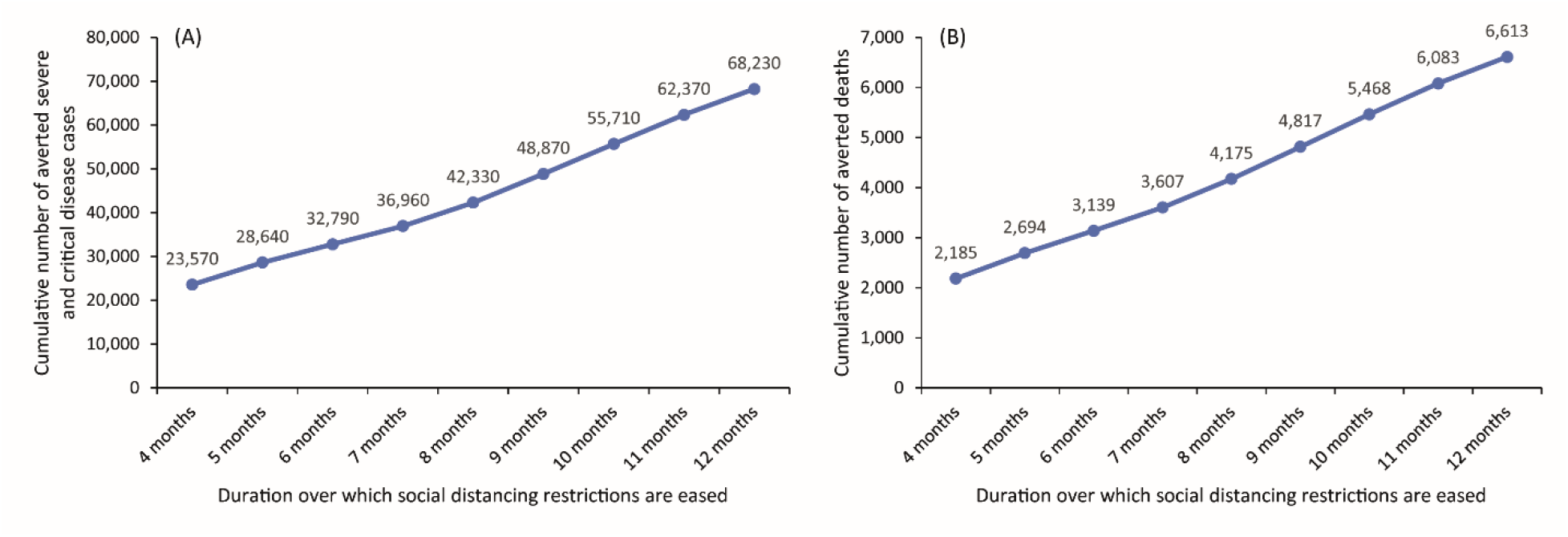
Impact of the duration of easing social and physical distancing restrictions on the cumulative number of averted (A) severe and critical disease cases and (B) deaths from 14 February 2021 (date of vaccine introduction) to December 31, 2021. The scenarios assume gradual easing of all restrictions over four months starting 15 April 2021 and reaching 80% vaccine coverage by 31 December 2021.

The extent of transmissibility of the variants of concern introduced on April 15 2021 remains uncertain. Analysis factoring in the uncertainty around the assumed R_0_ of these variants indicates that the value of R_0_ will affect the scale of the predicted surge (Figure S1A) and, hence, will affect the cumulative number of averted infections (Figure S1B), with higher R_0_ leading to more averted infections. However, the beneficial vaccine impact relative to the no vaccination scenario remains largely unchanged.

## Discussion

Our analysis predicts that current held immunity levels in Lebanon, estimated by the model at 40% on 15 April 2021, are still not sufficient to go back to normalcy and will result in a large new epidemic surge if variants of concern are introduced and if restrictions are eased. We found that for vaccination to make an impact on this predicted surge, its scale-up has to be rapid; otherwise, a large fraction of people in the community will be infected before they get vaccinated. For substantial impact, high levels of vaccine coverage also need to be achieved, as the gains from vaccination did not increase linearly with coverage and were most pronounced starting at 70% coverage. While beneficial, intermediate coverage will not be sufficient to allow return of social contacts to normalcy in the near term and will not spare substantial covid-19 hospitalizations and deaths. Reaching, for example, 40% coverage by the end of the year - which is the more likely scenario in Lebanon given current vaccination pace - means a roll-out that is too slow against fast moving incidence, resulting in a more limited impact of vaccination. Our analyses also highlight the importance of keeping social distancing restrictions as long as possible during vaccine scale-up to maximize the benefits of vaccination.

Therefore, vaccination will flatten the epidemic curve of a predicted summer surge in Lebanon but will not prevent it or delay it, unless scale-up to reach high vaccination coverage (at least 70%) will be very rapid – for example achieved in August rather than December 2021, while maintaining social distancing restriction during roll-out (Figure S2). At the pace vaccination is taking place, where only 3.3% of the population have completed two doses of the vaccine in three months, it is unlikely that vaccination will have a noticeable impact on a potential surge in the near future. The likelihood of this surge is high given the circulation worldwide of highly infectious SARS-CoV-2 variants of concern [13, 17, 38], the poor borders control implemented in the country, and absence of systematic viral genome sequencing or multiplex PCR variant screening to track the introduction of variants. Social contacts are also expected to increase over the summer months with the arrival of large numbers of Lebanese expatriates and other tourists. This is further motivated by a false sense of security due to the recent marked decline in the number of confirmed cases, as immunity had been building up mainly because of ongoing incidence [1]. There is therefore an urgent need to rapidly scale-up vaccination to avoid a surge that the country, with its multiple crises and limited health-care capacity, is largely unprepared for.

Multiple strategies can be followed to overcome supply, administration, and demand challenges to speed up community immunization in Lebanon. While some of the supply bottlenecks result from the high global demand on manufacturers and the prioritization of high-income countries for delivery of reserved doses due to early agreements [39], the government may adopt simple strategies to maximize the use of available doses. The recent decision by the Lebanese authorities to delay administration of the second dose is one good example [40]. Promoting the WHO-approved extraction of one additional jab per vial of the BNT162b2 vaccine [41, 42] and reinforcing prioritization of available doses for individuals at higher risk of developing severe disease [18, 35] are other suggestions that would capitalize on current supply and maximize its impact. Efforts also need to be put to expand the vaccine distribution campaign to cover all geographical and remote areas, and reach out to the most underprivileged communities. Currently, vaccination is being deployed only through major hospitals. Expanding vaccination sites to include primary health care centers and other existing health care infrastructures, as well as specifically establishing sites such as drive-through vaccination facilities or vaccination centers in sports arenas could be considered. Particular consideration needs to be made to reach the most vulnerable, including refugees, migrant workers, and other underprivileged communities who may not have access to the digital registration platform or to vaccination sites. Mobile vaccination units, partnering with municipalities, and engaging local community organizations would help enhance equitable distribution of the vaccine. Finally, the vaccination campaign should be accompanied by a strategic communication plan, clear and tailored awareness campaigns, and community engagement initiatives to address vaccine hesitancy and increasing the public’s confidence in vaccines and in the national vaccination program.

This study has some limitations. Model estimations are contingent on the validity and generalizability of input data. While current available natural history and epidemiological evidence was used to justify model assumptions and parameter values, our understanding of this infection is still evolving. Data on SARS-CoV-2 seroprevalence in the Lebanese population are lacking. The level of prior exposure to the infection plays an important role in this analysis and will affect estimates of vaccine impact. We assumed that 20% had been exposed to the infection on January 1,2021, a sensible estimate based on triangulation of available local data, including the cumulative number of confirmed infections and deaths and routine clinical antibody testing data. We assumed that both natural and vaccine-induced immunity last for one year, as suggested by available data [21-24]. However, these two important parameters remain unknown. If they prove to be less than the assumed one year, the impact of the vaccine will be reduced. The model did not prioritize vaccination based on age, since younger individuals in the workforce are currently being vaccinated through the private sector. Given that the highest risk elderly population (>70 years) did receive the vaccine first, our estimates for the impact of vaccination are conservative and the number of averted disease cases and deaths may be higher than the ones reported in our study. Finally, analyses were conducted at a national level, whereas vaccine impact will likely differ between geographic locations with different epidemic transmission dynamics, different levels of compliance to preventive measures, and different vaccine uptake levels.

In conclusion, amidst all ongoing crises in the country, Lebanon cannot afford the burden of yet another large epidemic wave. Given the dire economic situation, strict lockdowns and extended closures may also be difficult to sustain. At this stage, scaling up vaccination offers the only hope to go back to normalcy with the least deaths and burden on the health care system possible, and with the least damage to the economy in the short and longer term. Our findings reiterate that there is every virtue in speeding up vaccine roll-out and increasing coverage to avoid the potentially devastating consequences of a predicted and likely epidemic surge. With only 6.8 million inhabitants, this goal should be feasible. However, concerted efforts need to be put to overcome local challenges. In the meantime, public messages need to promote adherence to social distancing measures until we approach high levels of vaccine coverage.

## Data Availability

All data are available in the manuscript and its supplementary materials

## Supplementary materials

Text S1A: Model structure

Text S1B: Model equations

Text S1C: Model parameters

Table S1: Definitions of population variables and symbols used in the model

Table S2. Model assumptions in terms of parameter values

Figure S1: Analysis of the effect of uncertainty around the transmissibility (R0) of the new variants introduced on 15 April 2021

Figure S2: Impact of SARS-CoV-2 vaccination on number of A) new infections, B) new severe and critical disease cases, and C) new deaths in Lebanon for vaccine coverage of 80% by 31 August 2021

## Author Contributions

GRM co-conceptualized the study, co-designed the methodology, and wrote the first draft of the manuscript. MM constructed and parameterized the mathematical model, and conducted the mathematical modeling analyses. FE-J co-conceptualized the study. LJA-R co-conceptualized the study and led the design of the methodology. All authors contributed to the discussion and interpretation of the results, and writing of the manuscript. All authors have read and approved the final manuscript.

## Institutional Review Board Statement

Not applicable

## Informed Consent Statement

Not applicable.

## Data Availability Statement

All data are available in the manuscript and its supplementary materials.

## Acknowledgments

The authors are grateful for support provided by the Biomedical Research Program and the Biostatistics, Epidemiology, and Biomathematics Research Core, both at Weill Cornell Medicine-Qatar.

