## Supplemental Material for "Modeling the impact of COVID-19 vaccination in Lebanon: A call to speed-up vaccine roll out"

**Text S1. SARS-CoV-2 vaccine mathematical model**

**A. Model structure**

We extended a recently-developed age-structured deterministic compartmental model [1-9]. The model is illustrated in Figure S1. The model stratifies the unvaccinated and vaccinated populations into compartments according to age group (0-9, 10-19, 20-29,…, ≥80 years), infection status (uninfected, infected), infection stage (mild, severe, critical), disease stage (severe, critical), and compartments for the gamma distribution (Γ-distribution) describing the waning of natural and vaccine immunity.

Transmission and disease progression dynamics in the vaccinated and unvaccinated cohorts are described in the model using age-specific sets of nonlinear ordinary differential equations, where each age group $a$ ($a = 1, 2, ...9$) refers to a 10-year age band (0-9,10-19,..70-79) apart from the last group including all those aged ≥0 years. The model is illustrated in Figure S1.

**Figure S1. Schematic diagram describing the SARS-CoV-2 transmission dynamics model in presence of a vaccine that reduces susceptibility to infection.**

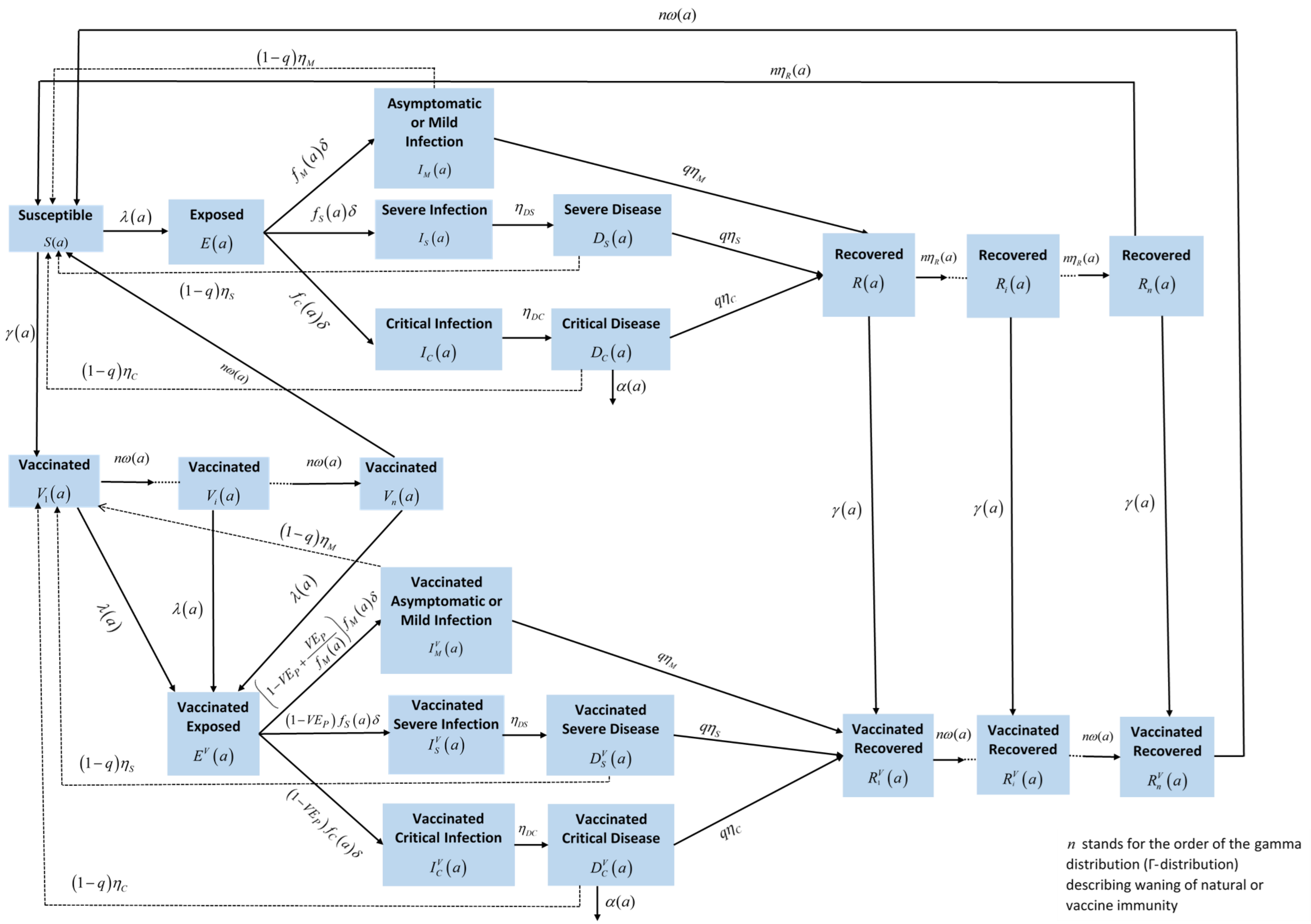

### B. Model equations

The definitions of population variables and symbols used in the equations are in Table S1below.

Unvaccinated population:

$$\frac{dS(a)}{dt} = \xi(a-1)S(a-1) - \left(\lambda(a) + \mu + \xi(a) + \gamma(a)\right)S(a) + (1-q)\left(\eta_M I_M(a) + \eta_S I_S(a) + \eta_C I_C(a)\right)$$
$$+ n\omega(a)\left(V_n(a) + R_n^V(a)\right) + n\eta_R R_n(a)$$

$$\frac{dE(a)}{dt} = \xi(a-1)E(a-1) + \lambda(a)S(a) - \left(\delta + \mu + \xi(a)\right)E(a)$$

$$\frac{dI_M(a)}{dt} = \xi(a-1)I_M(a-1) + f_M(a)\delta E(a) - \left(\eta_M + \mu + \xi(a)\right)I_M(a)$$

$$\frac{dI_S(a)}{dt} = \xi(a-1)I_S(a-1) + f_S(a)\delta E(a) - \left(\eta_{DS} + \mu + \xi(a)\right)I_S(a)$$

$$\frac{dI_C(a)}{dt} = \xi(a-1)I_C(a-1) + f_C(a)\delta E(a) - \left(\eta_{DC} + \mu + \xi(a)\right)I_C(a)$$

$$\frac{dD_S(a)}{dt} = \xi(a-1)D_S(a-1) + \eta_{DS} I_S(a) - \left(\eta_S + \mu + \xi(a)\right)D_S(a)$$

$$\frac{dD_C(a)}{dt} = \xi(a-1)D_C(a-1) + \eta_{DC} I_C(a) - \left(\eta_C + \mu + \xi(a) + \alpha(a)\right)D_C(a)$$

$$\frac{dR_1(a)}{dt} = \xi(a-1)R_1(a-1) + \left(q\eta_M I_M(a) + q\eta_S D_S(a) + q\eta_C D_C(a)\right) - \left(\mu + \xi(a) + \gamma(a) + n\eta_R\right)R_1(a)$$

For k=2,..n

$$\frac{dR_k(a)}{dt} = \xi(a-1)R_k(a-1) + n\eta_R R_{k-1}(a) - \left(\mu + \xi(a) + \gamma(a) + n\eta_R\right)R_k(a)$$

Vaccinated populations aged 10+ years:

$$\frac{dV_1(a)}{dt} = \xi(a-1)V_1(a-1) + \gamma(a)S(a) + (1-q)\left(\eta_M I_M^V(a) + \eta_S I_S^V(a) + \eta_C I_C^V(a)\right)$$
$$-\left((1-VE_S)\lambda(a) + \mu + \xi(a) + n\omega(a)\right)V_1(a)$$

For k=2,..n

$$\frac{dV_k(a)}{dt} = \xi(a-1)V_k(a-1) + n\omega(a)V_{k-1}(a) - \left((1-VE_S)\lambda(a) + \mu + \xi(a) + n\omega(a)\right)V_k(a)$$

$$\frac{dE^V(a)}{dt} = \xi(a-1)E^V(a-1) + (1-VE_S)\lambda(a)\left(\sum_{k=1}^{n} V_k(a)\right) - (\delta + \mu + \xi(a))E^V(a)$$

$$\frac{dI_M^V(a)}{dt} = \xi(a-1)I_M^V(a-1) + f_M(a)\delta E^V(a) - (\eta_M + \mu + \xi(a))I_M^V(a)$$

$$\frac{dI_S^V(a)}{dt} = \xi(a-1)I_S^V(a-1) + f_S(a)\delta E^V(a) - (\eta_{DS} + \mu + \xi(a))I_S^V(a)$$

$$\frac{dI_C^V(a)}{dt} = \xi(a-1)I_C^V(a-1) + f_C(a)\delta E^V(a) - (\eta_{DC} + \mu + \xi(a))I_C^V(a)$$

$$\frac{dD_S^V(a)}{dt} = \xi(a-1)D_S^V(a-1) + \eta_{DS}I_S^V(a) - (\eta_S + \mu + \xi(a))D_S^V(a)$$

$$\frac{dD_C^V(a)}{dt} = \xi(a-1)D_C^V(a-1) + \eta_{DC}I_C^V(a) - (\eta_C + \mu + \xi(a) + \alpha(a))D_C^V(a)$$

$$\frac{dR_1^V(a)}{dt} = \xi(a-1)R_1^V(a-1) + \gamma(a)R_1(a) + q\eta_M I_M^V(a) + q\eta_S D_S^V(a) + q\eta_C D_C^V(a) - (\mu + \xi(a) + n\omega(a))R_1^V(a)$$

For k=2,..n

$$\frac{dR_k^V(a)}{dt} = \xi(a-1)R_k^V(a-1) + \gamma(a)R_k(a) + n\omega(a)R_{k-1}^V(a) - (\mu + \xi(a) + n\omega(a))R_k^V(a)$$

Where $n$ stands for the order of the gamma distribution (Γ-distribution) describing waning of natural or vaccine immunity.

**Table S1. Definitions of population variables and symbols used in the model.**

| Symbol | Definition |
|---|---|
| **Transmission dynamics parameters** | |
| $S(a)$ | Unvaccinated susceptible population |
| $E(a)$ | Unvaccinated latently infected population |
| $I_M(a)$ | Unvaccinated population with asymptomatic or mild infection |
| $I_S(a)$ | Unvaccinated population with severe infection |
| $I_C(a)$ | Unvaccinated population with critical infection |
| $D_S(a)$ | Unvaccinated population with severe disease |
| $D_C(a)$ | Unvaccinated population with critical disease |
| $R_i(a)$ | $i^{th}$ compartment to generate the gamma distribution for the waning of natural immunity among the unvaccinated recovered population |

| Symbol | Definition |
|---|---|
| $V_i(a)$ | $i^{th}$ compartment to generate the gamma distribution for the waning of vaccine immunity among the vaccinated susceptible population |
| $E^V(a)$ | Vaccinated latently infected population |
| $I_M^V(a)$ | Vaccinated population with asymptomatic or mild infection |
| $I_S^V(a)$ | Vaccinated population with severe infection |
| $I_C^V(a)$ | Vaccinated population with critical infection |
| $D_S^V(a)$ | Vaccinated population with severe disease |
| $D_C^V(a)$ | Vaccinated population with critical disease |
| $R_i^V(a)$ | $i^{th}$ compartment to generate the gamma distribution for the waning of natural immunity among the vaccinated recovered population |
| $N$ | Total population size |
| $n_{age}$ | Number of age groups |
| $n$ | Order of the gamma distribution (Γ-distribution) describing waning of natural or vaccine immunity |
| $\xi(a)$ | Transition rate from one age group to the next age group |
| $\beta$ | Overall infectious contact rate |
| $1/\delta$ | Duration of latent infection |
| $1/\eta_M$ | Duration of asymptomatic or mild infection |
| $1/\eta_{DS}$ | Duration of severe infection infectiousness before isolation and/or hospitalization |
| $1/\eta_S$ | Duration of severe disease following onset of severe disease |
| $1/\eta_{DC}$ | Duration of critical infection infectiousness before isolation and/or hospitalization |
| $1/\eta_C$ | Duration of critical disease following onset of critical disease |
| $1/\eta_R$ | Duration of natural immunity |
| $1/\mu$ | Natural death rate |
| $\alpha(a)$ | Mortality rate in each age group |
| $f_M(a)$ | Proportion of infections that will progress to be mild or asymptomatic infections |
| $f_S(a)$ | Proportion of infections that will progress to be severe infections |
| $f_C(a)$ | Proportion of infections that will progress to be critical infections |
| $q$ | Proportion of infected people who do not develop natural immunity and become again susceptible upon recovery |
| **Key vaccine product characteristics** | |
| $VE_s$ | Vaccine efficacy in reducing susceptibility |
| $1/\omega$ | Duration of vaccine protection |

The force of infection (hazard rate of infection) experienced by the unvaccinated susceptible populations $S(a)$ is given by

$$\lambda(a)=\beta\sum_{a'=1}^{n_{age}}\mathcal{H}_{a,a'}\frac{I_M(a')+I_S(a')+I_C(a')+I_M^V(a')+I_S^V(a')+I_C^V(a')}{\left[\begin{array}{l}S(a')+E(a')+I_M(a')+I_S(a')+I_C(a')+D_S(a')+D_C(a')+\sum_{i=1}^{n}R_i(a')+\\\sum_{i=1}^{n}V_i(a')+E^V(a')+I_M^V(a')+I_S^V(a')+I_C^V(a')+D_S^V(a')+D_C^V(a')+\sum_{i=1}^{n}R_i^V(a')\end{array}\right]},$$

while that experienced by the vaccinated susceptible populations $V(a)$ is given by

$$\lambda^V(a)=(1-VE_S)\lambda(a)$$

where $\beta$ is the overall infectious contact rate, parametrized using a combination of Wood-Saxon and logistic functions:

$$\beta(t)=\underbrace{\frac{a_1}{1+e^{\left(\frac{t-b_1}{c_1}\right)}}}_{Wood\text{-}Saxon\ function}+\underbrace{\frac{a_2}{1+e^{-\left(\frac{t-b_2}{c_2}\right)}}}_{Logistic\ function}$$

A temporal variation in the infectious contact rate was considered to accommodate the time evolution of SARS-CoV-2 epidemic in Lebanon. Here $a_1$, $a_2$, $b_1$, $b_2$, $c_1$, and $c_2$ are fitting parameters.

The mixing among the different age groups is dictated by the mixing matrix $\mathcal{H}_{a,a''}$. This matrix provides the probability that an individual in the $a$ age group will mix with an individual in the $a'$ age group (regardless of vaccination status). The mixing matrix is given by

$$\mathcal{H}_{a,a'}=e_{Age}\delta_{a,a'}+(1-e_{Age})\frac{\left[\begin{array}{l}S(a')+E(a')+I_M(a')+I_S(a')+I_C(a')+D_S(a')+D_C(a')+\sum_{i=1}^{n}R_i^V(a')+\\\sum_{i=1}^{n}V_i(a')+E^V(a')+I_M^V(a')+I_S^V(a')+I_C^V(a')+D_S^V(a')+D_C^V(a')+\sum_{i=1}^{n}R_i^V(a')\end{array}\right]}{\sum_{a=1}^{n_{age}}\left[\begin{array}{l}S(a)+E(a)+I_M(a)+I_S(a)+I_C(a)+D_S(a)+D_C(a)+\sum_{i=1}^{n}R_i^V(a)+\\\sum_{i=1}^{n}V_i(a)+E^V(a)+I_M^V(a)+I_S^V(a)+I_C^V(a)+D_S^V(a)+D_C^V(a)+\sum_{i=1}^{n}R_i^V(a)\end{array}\right]}$$

Here, $\delta_{a,a'}$ is the identity matrix. $e_{Age}\in[0,1]$ measures the degree of assortativeness in the mixing. At the extreme $e_{Age}=0$, the mixing is fully proportional. Meanwhile, at the other extreme, $e_{Age} = 1$, the mixing is fully assortative, that is individuals mix only with members in their own age group.

### C. Parameter values

The input parameters of the model were chosen based on current empirical data for SARS-CoV-2 natural history and epidemiology. The parameter values are listed in Table S2.

**Table S2. Model assumptions in terms of parameter values**

| Parameter | Symbol | Value | Justification |
|---|---|---|---|
| Duration of latent infection | $1/\delta$ | 3.69 days | Based on existing estimate [10] and based on a median incubation period of 5.1 days [11] adjusted by observed viral load among infected persons [12] and reported transmission before onset of symptoms [13] |
| Duration of infectiousness | $1/\eta_M$ ; $1/\eta_{DS}$ ; $1/\eta_{DC}$ | 3.48 days | Based on existing estimate [10] and based on observed time to recovery among persons with mild infection [10, 14] and observed viral load in infected persons [12, 13, 15] |
| Duration of severe disease following onset of severe disease | $1/\eta_S$ | 28 days | Observed duration from onset of severe disease to recovery [14] |
| Duration of hospitalization for critical infection | $1/\eta_C$ | 42 days | Observed duration from onset of critical disease to recovery [14] |
| Life expectancy | $1/\mu$ | 79.10 years | United Nations World Population Prospects database [16] |
| Proportion of infections that will progress to be mild or asymptomatic infections | $f_M(a)$ | Determined from $f_M(a)+f_S(a)+f_C(a)=1$ | Observed proportion of infections that eventually develop mild or asymptomatic in France [14, 17, 18] |
| Proportion of infections that will progress to be infections that require hospitalization in acute care beds | $f_S(a)$ | | The distribution and age dependence of asymptomatic/mild, severe, or critical infections was based on the modeled SARS-CoV-2 epidemic in France [19] |
| Age 0-19 years | $RRS1\times f_S$ | $RRS1=0.1$ | Model-estimated relative risk of severe infection based on the SARS-CoV-2 epidemic in France [19] |
| Age 20-29 years | $RRS2\times f_S$ | $RRS2=0.5$ | Model-estimated relative risk of severe infection based on the SARS-CoV-2 epidemic in France [19] |
| Age 30-39 years | $f_S$ | Reference category ( $f_S$ = 0.01) | Model-estimated based on fitting the SARS-CoV-2 epidemic in France [19] |

| | | | |
|---|---|---|---|
| Age 40-49 years | $RRS3 \times f_S$ | $RRS3 = 1.2$ | Model-estimated relative risk of severe infection based on the SARS-CoV-2 epidemic in France [19] |
| Age 50-59 years | $RRS4 \times f_S$ | $RRS4 = 2.3$ | Model-estimated relative risk of severe infection based on the SARS-CoV-2 epidemic in France [19] |
| Age 60-69 years | $RRS5 \times f_S$ | $RRS5 = 4.5$ | Model-estimated relative risk of severe infection based on the SARS-CoV-2 epidemic in France [19] |
| Age 70-79 years | $RRS6 \times f_S$ | $RRS6 = 7.8$ | Model-estimated relative risk of severe infection based on the SARS-CoV-2 epidemic in France [19] |
| Age 80+ years | $RRS7 \times f_S$ | $RRS7 = 27.6$ | Model-estimated relative risk of severe infection based on the SARS-CoV-2 epidemic in France [19] |
| Proportion of infections that will progress to be critical infections | $f_C(a)$ | | The distribution and age dependence of asymptomatic/mild, severe, or critical infections was based on the modeled SARS-CoV-2 epidemic in France [19] |
| Age 0-19 years | $RRC1 \times f_C$ | $RRC1 = 0.21$ | Model-estimated relative risk of critical infection based on the SARS-CoV-2 epidemic in France [19] |
| Age 20-29 years | $RRC2 \times f_C$ | $RRC2 = 0.33$ | Model-estimated relative risk of critical infection based on the SARS-CoV-2 epidemic in France [19] |
| Age 30-39 years | $f_C$ | Reference category $f_c = 0.0002$ | Model-estimated based on fitting the SARS-CoV-2 epidemic in France [19] |
| Age 40-49 years | $RRC3 \times f_C$ | $RRC3 = 1.83$ | Model-estimated relative risk of critical infection based on the SARS-CoV-2 epidemic in France [19] |
| Age 50-59 years | $RRC4 \times f_C$ | $RRC4 = 4.67$ | Model-estimated relative risk of critical infection based on the SARS-CoV-2 epidemic in France [19] |
| Age 50-59 years | $RRC4 \times f_C$ | $RRC4 = 4.67$ | Model-estimated relative risk of critical infection based on the SARS-CoV-2 epidemic in France [19] |
| Age 60-69 years | $RRC5 \times f_C$ | $RRC5 = 10.58$ | Model-estimated relative risk of critical infection based on the SARS-CoV-2 epidemic in France [19] |
| Age 70-79 years | $RRC6 \times f_C$ | $RRC6 = 13.61$ | Model-estimated relative risk of critical infection based on the SARS-CoV-2 epidemic in France [19] |
| Age 80+ years | $RRC7 \times f_C$ | $RRC7 = 8.67$ | Model-estimated relative risk of critical infection based on the SARS-CoV-2 epidemic in France [19] |
| Disease mortality rate in each age group | $\alpha(a)$ | | The distribution and age dependence of COVID-19 mortality was based on the modeled SARS-CoV-2 epidemic in France [19] |
| Age 0-19 years | $RRD1 \times \alpha$ | $RRD1 = 0.10$ | Model-estimated relative risk of death based on the SARS-CoV-2 epidemic in France [19] |
| Age 20-29 years | $RRD2 \times \alpha$ | $RRD2 = 0.40$ | Model-estimated relative risk of death based on the SARS-CoV-2 epidemic in France [19] |

| | | | |
|---|---|---|---|
| Age 30-39 years | $\alpha$ | Reference category ( $\alpha$ =0.00096) | Model-estimated based on fitting the number of SARS-CoV-2 deaths in Lebanon [20] |
| Age 40-49 years | $RRD3 \times \alpha$ | $RRD3 = 3.00$ | Model-estimated relative risk of death based on the SARS-CoV-2 epidemic in France [19] |
| Age 50-59 years | $RRD4 \times \alpha$ | $RRD4 = 10.00$ | Model-estimated relative risk of death based on the SARS-CoV-2 epidemic in France [19] |
| Age 60-69 years | $RRD5 \times \alpha$ | $RRD5 = 45.00$ | Model-estimated relative risk of death based on the SARS-CoV-2 epidemic in France [19] |
| Age 70-79 years | $RRD6 \times \alpha$ | $RRD6 = 120.00$ | Model-estimated relative risk of death based on the SARS-CoV-2 epidemic in France [19] |
| Age 80+ years | $RRD7 \times \alpha$ | $RRD7 = 505.00$ | Model-estimated relative risk of death based on the SARS-CoV-2 epidemic in France [19] |

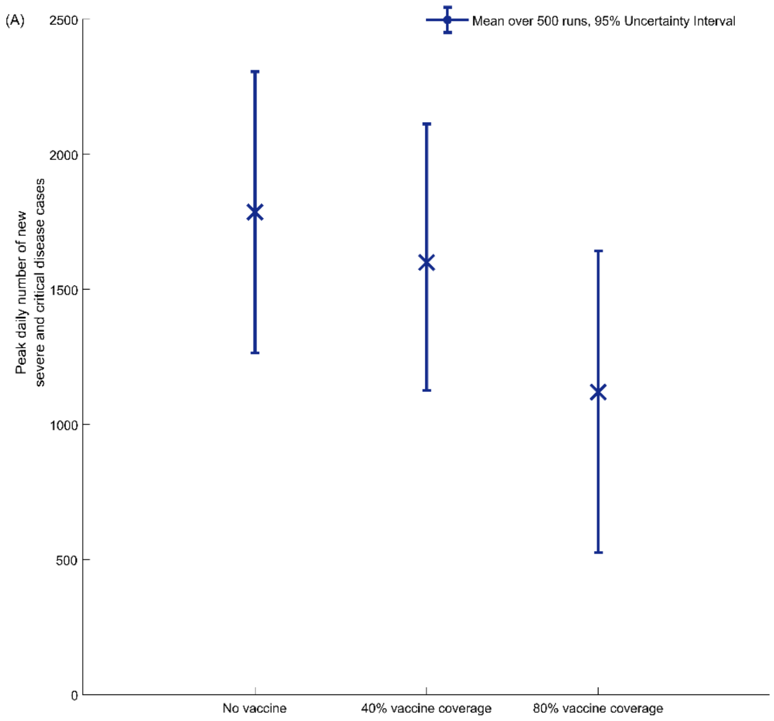


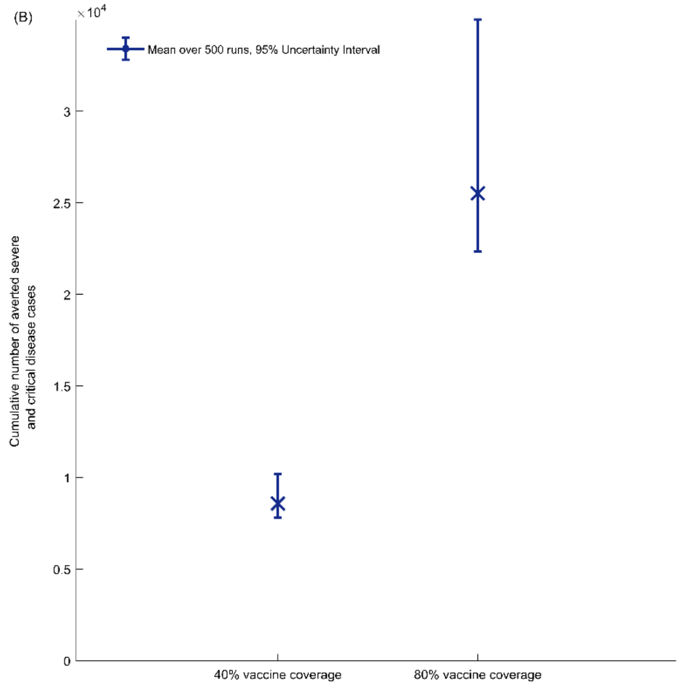


**Figure S1.** Analysis of the effect of uncertainty around the transmissibility ($R_0$) of the new variants introduced on 15 April 2021. Mean over 500 runs and 95% uncertainty intervals for (A) the peak daily number of new severe/disease cases and (B) the cumulative number of averted severe/disease cases between 14 February 2020 (day of vaccine introduction) and 31 December 2021, for the two scenarios of 40% and 80% vaccine coverage by end of 2021.

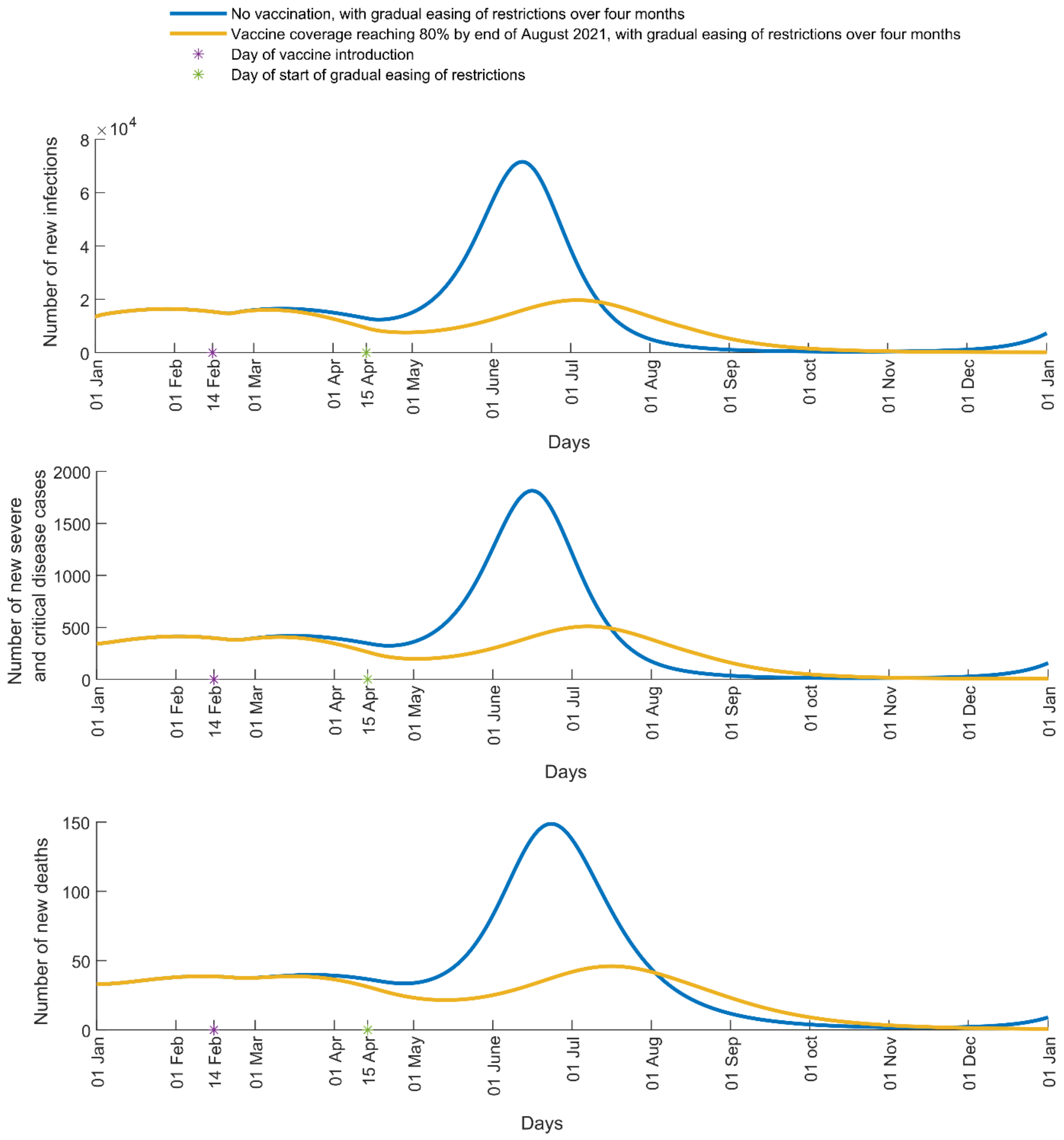


**Figure S2.** Impact of SARS-CoV-2 vaccination on number of A) new infections, B) new severe and critical disease cases, and C) new deaths in Lebanon. The vaccine is introduced on 14 February 2021 and vaccine coverage is scaled up to reach 80% (yellow curve) by 31 August 2021. The simulations assume $R_0$ of 1.2 from 1 January 2021 to April 15 2021 when it starts to increase with gradual easing of restrictions to reach 6.0 after four months.
